# Follow-up after post-exposure prophylaxis before and during the COVID-19 pandemic in Brazil

**DOI:** 10.1101/2024.03.07.24303803

**Authors:** Elaine Monteiro Matsuda, Ivana Barros de Campos, Luís Fernando de Macedo Brígido

## Abstract

Although post-exposure prophylaxis (PEP) is a powerful tool to abort HIV infection within 72 hours of exposure, blocking the establishment of chronic infection, follow-up metrics of this intervention are scarce. As antiretroviral use delays diagnosis biomarkers, the moment to perform serological evaluations must be considered this to avoid missed diagnosis opportunities. We assessed the return adherence after PEP dispensation in service in the Sao Paulo metropolitan area and reviewed the literature, both showing limited adherence to current protocols and leading to difficulties in diagnosing early HIV infection. The current proposed date for the first return after PEP is associated with low adherence and may have limited capability to detect antibodies if the infection is present. Guidelines should allow a longer time after PEP discontinuation along with message reminders to encourage adherence and avoid false negative results that can be detrimental both to the patient and to the community.

## INTRODUCTION

The Joint United Nations Programme on HIV/AIDS (UNAIDS) leads and inspires the world to achieve its shared vision of zero new HIV infections, zero discrimination, and zero AIDS-related deaths. Since 2010, new HIV infections have declined by 32%, from 2.2 million to 1.5 million in 2021^1^.

Combination prevention programs include a mix of evidence-based biomedical, behavioral, and structural interventions to meet the current HIV prevention needs of individuals and communities, aiming for the greatest possible impact on reducing the number of people newly infected^2^. They must be appropriate to each individual’s circumstances and HIV vulnerability^3^. Globally, gay men and other men who have sex with men are 28 times more likely to be infected with HIV. People who inject drugs have 35 times the risk, sex workers 30 times, and transgender women 14 times the risk^1^.

Antiretrovirals can provide not only treatment but also act as a preventive intervention through viral suppression that makes the individual undetectable = untransmissible^4^. Moreover, antiretroviral has been shown to be effective in pre-exposure prophylaxis (PrEP)^5^ and post-exposure (PEP)^6,7^ and is part of the main core of strategies for controlling the HIV epidemic^8^. The preferred regimen to the first line of treatment in Brazil is the same as that used for PEP and consists of tenofovir 300mg/lamivudine 300mg (TDF/3TC) associated with dolutegravir 50mg (DTG) daily^9^.

Brazilian as well as other guidelines recommend PEP with 3 drugs, prescribed after a point-of-care serological HIV test and dispensed for 28 days. PEP is recommended only within 72 hours of exposure, with guidance to repeat the HIV test^9,10,11,12,13^. The timing of this follow-up testing varies between four to six weeks and 12 weeks after exposure^9,10^, at the end of PEP and 10 to 12 weeks after exposure^12^, at a minimum of 45 days after completion of the PEP course, if the 28-day PEP course is completed, this is 73 days (10.5 weeks) post exposure^11^, and at 3 months after exposure^13^. CDC (USA) and the UK recommend the use of a fourth-generation test at the beginning of PEP, and if not used, the CDC recommends an additional serological follow-up 6 months after exposure^10,11^. The seroreactivity of the rapid test depends on the sensitivity of the test in relation to previous exposures (immunological window). The fourth-generation rapid test is more efficient in detecting very recent infections, even detecting antibodies not detected in the third-generation rapid test, as well as acute infection with the detection of the p24 antigen^14^.

The efficacy of PEP depends on the timing and proper use of the regimen. Delayed initiation of PEP, poor/non-adherence to the regimen, especially in the first days, and further high-risk sexual exposures after cessation of PEP may compromise the outcome. Moreover, early/primary HIV infection already established at the time of PEP initiation is a possibility in many situations^11^. Diagnosis of acute/early HIV infection, proper adherence to PEP protocols, as well as, laboratory follow-up are constant challenges to this policy^9,10,11,15,16^.

To evaluate the issue of post-PEP serological monitoring we carried out this study in a reference service that cares for people living with HIV and provides antiretroviral prophylaxis, PEP, and PrEP, to those who seek it spontaneously or were referred from other services, in Santo André, a metropolitan area of São Paulo/Brazil.

## METHODOLOGY

The Medication Logistic Control System (SICLOM) provided information on users with PEP dispensation between 2019 and 2021. Medical records were consulted in order to assess adherence to the recommended 30 and 120-day returns after risk exposure and other variables such as sex (female or male), gender (cis or transgender), men who have sex with men (MSM), sex worker (yes or not) and category of risk exposure (biological material exposition, occupational or not, sexual consent or not and others). Return after starting PEP between 26 and 40 days was considered for this study as a 30-day return and between 110 and 130 days as a 120-day return. Return on any date within 180 days was also evaluated.

Data obtained from electronic databases were anonymized before analysis. Statistical analyzes were performed with Stata version 14.2 (Stata Corp LLC, College Station, Texas, USA) and IBM SPSS Statistics for Windows, Version 24.0. (Armonk, NY, USA). The age (years) was expressed in medians, with the 25th and 75th percentiles (IQR). A significant level of p<0.05, two-tailed, was applied to all analyses. Variables were compared using Mann-Whitney or Kruskal-Wallis test for continuous variables and chi-squared (χ2) or Fisher’s exact tests for categorical variables, as appropriate.

## RESULTS

During the study period, we obtained 2168 PEP events recorded at SICLOM, dispensed for 1468 users. Additional information could be obtained only from 1281/1468 users. The median age of these users was 31 years (IQR25-75 24-39), with 6/1281 0.3% being under 14 years and 17/1281 0.8% above 60 years.

Table 1 describes demographic characteristics by year of study. Most were male (853/1281 67%), with 368/853 43% of this reporting being MSM, 39/853 4.6% identified as transgender women (TW), which corresponds to 27/931 2.9% among all users. Almost all TW were sex workers, 90% 35/39 versus 2.4% 29/1207 among ciswomen (p<0.0001). Among cisgender, the proportion of sex workers among women was higher than among men, 5.4% 23/428 versus 0.7% 6/808 (p<0.0001).

**Table 1.** Demographic characteristics among cases with the dispensation of post-exposure prophylaxis in each year of the study.

|  | ALL | 2019 | 2020 | 2021 | p |
| --- | --- | --- | --- | --- | --- |
| N | 1281 | 636 (49.6%) | 422 (32.9%) | 223 (17.4%) | 1 |
| Age (n years) | 31<br>(IQR25-75 24-39) | 30<br>(IQR 23-38) | 31<br>(IQR 25-38) | 32<br>(IQR 25-41) | 0.02 |
| Female | 428/1281<br>33.4% | 199/636<br>31.3% | 116/422<br>27.5% | 113/223<br>50.7% | 0.0001 |
| Male | 853/1281<br>66.6% | 437/636<br>68.7% | 306/422<br>72.5% | 110/223<br>49.3% |  |
| MSM | 368/853<br>43.1% | 198/437<br>45,3% | 127/306<br>41,5% | 43/110<br>39,1% | 0.36 |
| Transwoman | 39/853<br>4.6% | 20/437<br>4,58% | 13/306<br>4,25% | 6/110<br>5,46% | 0.87 |
| Sex workers | 64/1275<br>5% | 42/631<br>6.7% | 17/422<br>4% | 5/222<br>2.3% | 0.02 |

MSM, man who have sex with man

We verified a change in the profile of PEP users who sought the service, still young adults, but with increasing age, with a median of 30, 31, and 32 years, in 2019, 2020, and 2021, respectively (p=0.02) and a proportional increase of women 31%, 28% and 51% (p<0.0001), which may be due in part to the increase of occupational accidents during the study period 27%, 33% and 53%, mostly women 70%, 74%, 76%.

Information regarding the category of risk exposure that motivated the search for PEP referred to in the medical records and in which group (female sex, MSM, TW, and/or sex worker) are summarized in Table 2.

**Table 2.** Type of risk exposure for HIV infection that motivated the dispensation of post-exposure prophylaxis in the different risk exposure categories and in each year of the study.

|  | ALL<br>n=1276 | 2019<br>635/1281 (49.6%) | 2020<br>419/1281 (32.9%) | 2021<br>222/1281 (17.4%) |
| --- | --- | --- | --- | --- |
| Accident with biological material |  |  |  |  |
| Occupational | 389/1276 30.5%<br>283/389 72.8% female<br>0 MSM<br>0 TW<br>0 sex worker | 169/635 26.91%<br>118/169 69.8% female<br>0 MSM<br>0 TW<br>0 sex worker | 103/419 32.7%<br>76/103 73.8% female<br>0 MSM<br>0 TW<br>0 sex worker | 117/222 52.7%<br>89/117 76.1% female<br>0 MSM<br>0 TW<br>0 sex worker |
| Non-occupational | 12/1276 0.9%<br>6/12 50% female<br>2/6 33.4% male MSM<br>0 TW<br>0 sex worker | 6/635 0.9%<br>3/6 female 50%<br>1/3 33.4% male MSM<br>0 TW<br>0 sex worker | 5/419 1.2%<br>2/5 female 40%<br>1/3 33.4% male MSM<br>0 TW<br>0 sex worker | 1/222 0.5 %<br>1/1 female 100%<br>0 MSM<br>0 TW<br>0 sex worker |
| Sexual |  |  |  |  |
| Sexual Consent | 831/1276 65.1%<br>101/831 12% female<br>364/730 49.9% male MSM<br>39/730 5.3% male TW<br>64/830 7.7% sex worker | 436/635 68.7%<br>60/436 13.8% female<br>196/376 52.1% male MSM<br>20/376 5.3% male TW<br>42/436 9.6% sex worker | 300/419 71.6%<br>27/300 9% female<br>125/273 45.8% male MSM<br>13/273 4.8% male TW<br>17/300 5.7% sex worker | 95/222 42.8%<br>14/95 14.8% female<br>43/81 53% male MSM<br>6/81 7.4% male TW<br>5/94 5.3% sex worker |
| Sexual Assault | 38/1276 2.98%<br>36/38 94.7% female<br>1/2 50% male MSM<br>0 TW<br>0 sex worker | 18/635 2.83%<br>17/18 94.5% female<br>0 MSM<br>0 TW<br>0 sex worker | 11/419 2.6%<br>10/11 90.9% female<br>1/1 100% male MSM<br>0 TW<br>0 sex worker | 9/222 4.1%<br>9/9 100% female<br>0 MSM<br>0 TW<br>0 sex worker |
| Other | 6/1276 0.47 %<br>1/6 16.7% female<br>1/5 20% male MSM<br>0 TW<br>0 sex worker | 6/635 0.94%<br>1/6 16.7% female<br>1/5 20% male MSM<br>0 TW<br>0 sex worker | 0 | 0 |

Table 3 demonstrates adherence to returns of 30 and 120 days isolated and associated, and any time up to 180 days. There was a reduction in returns at any time after PEP during the COVID-19 pandemic, from 39.5% in 2019 to 12.8% (2020) and 20.2% (2021), (p<0.001). The adherence to the 30-day return was also smaller in the years 2020 and 2021 compared to the year 2019 (p=0.0001). However, from 2019 to 2021, if we analyze the 30-day versus 120-day returns separately, the adherence was greater in the 30-day return, 315/1281 24,6% versus 103/1281 8% (p<0.0001).

**Table 3.** Adherence to the 30 and 120-day returns, isolated and in different associations, or at any time within 180 days after exposure to risk for HIV infection.

| Adherence |  | ALL | 2019 | 2020 | 2021 | p |
| --- | --- | --- | --- | --- | --- | --- |
| 30-day return | **yes** | **315 24.6%** | **230 36.3%** | **45 10.7%** | **38 17%** | **0.0001** |
|  | no | 966 75.4% | 404 63.7% | 377 54.7% | 185 83% |  |
| Return between 30 and 120 days, median of 57days (IQR25-75 43-67) | **yes** | **79 6.2%** | **60 9.4%** | **9 2.1%** | **10 4.5%** | **0.12** |
|  | no | 1202 93.8% | 576 90.6% | 413 97.9% | 213 95.5% |  |
| 120-day return | **yes** | **103 8%** | **77 12.2%** | **13 3.1%** | **12 5.4%** | **0.03** |
|  | no | 1178 92% | 557 87.8% | 409 96.9% | 211 94.6% |  |
| Return of 30 and 120 days | **yes** | **65 5.1%** | **56 8.8%** | **4 0.9%** | **5 2.2%** | **0.07** |
|  | no | 1216 94.9% | 580 8.8% | 418 99.1% | 218 97.8% |  |
| Return 30 and absence 120 days | **yes** | **247 19.3%** | **173 27.2%** | **41 9.7%** | **33 14.8%** | **0.0001** |
|  | no | 1034 80.7% | 463 72.8% | 381 90.3% | 190 85.2% |  |
| Absence in 30 and return in 120 | **yes** | **37 2.9%** | **21 2.6%** | **9 2.1%** | **7 3.1%** | **0.95** |
|  | no | 1244 97.1% | 615 96.7% | 413 97.9% | 96.90% |  |
| Absence in 30 and return 120 days | **yes** | **931 72.7%** | **385 60.5%** | **368 87.2%** | **178 79.8%** | **0.0001** |
|  | no | 350 16.1% | 251 39.5% | 54 12.8% | 45 20.2% |  |
| Return at any time | yes | **350 27.3%** | **251 39.5%** | **54 12.8%** | **45 20.2%** | **<0.001** |
|  | no | 931 72.7% | 385 60.5% | 368 87.3% | 178 79.8% |  |

## DISCUSSION

PEP is an efficacious HIV prevention option that has been underutilized, representing a missed opportunity to prevent or abort HIV infection associated with high-risk exposures^10,17,18^.

Ruling out acute HIV infection prior to prophylactic antiretroviral use is particularly challenging in low-and middle-income settings, where there is limited access to advanced laboratory testing and infrastructure^15^. As the 3-drug PEP regimen is the same as that used in first-line treatment (tenofovir/lamivudine + dolutegravir), when the HIV infection is not blocked by PEP (viral infection is established), or starting PEP in a patient in the acute/early phase, both cases, will be on early therapy. This very early treatment has been suggested as potentially beneficial to the patient^19,20^ and avoids further viral transmission at this highly infectious phase^20,21^. However, recognition of infection is cumbersome at this stage, and several studies demonstrate a delay in seroconversion and viremia detection of HIV-1, due to the use of antiretroviral drugs, preventing proper use of serological and other biomarkers of infection^15,16^. This increases the probability of negative false results in HIV testing, allowing an undiagnosed patient to return to the community with an uncontrolled viremia. Better diagnosis approaches to this situation are clearly needed. This delay in seroconversion becomes even more worrying in cases where PrEP is prescribed, in which the two-drug scheme used in PrEP will be a sub-optimal treatment regimen that, as a consequence of an undocumented infection, implies the risk of inducing resistance mutations and virological failure^22^.

Manak et al. evaluated the performance of HIV antigen/antibody combination at weeks 12 and 24 following the initiation of antiretroviral therapy (ART) at Fiebig stage I (FI), FII, or FIII/IV in comparison to samples from untreated cases, who demonstrated robust reactivity, while 52.2% of samples from individuals initiating ART at FI, 7.7% at FII, and 4.5% at FIII/IV were nonreactive by the HIV Ag/Ab Combo assays^16^. Although the first evaluation in the use of ART was at 12 weeks, it would be expected that there would also be this delay with 4 weeks of the use PEP or PrEP.

While excellent, well-tolerated treatment regimens are available, adherence to PEP medications and attendance at clinical visits may be sub-optimal in certain groups of individuals^8^. In an Australian cohort of mainly MSM, only 34% of 1864 had follow-up testing at 12 weeks after initiation of PEP^23^. Several studies in the UK report that attendance at the 12-week follow-up HIV test is poor (30–67%)^11^.

In our service, the first year of the COVID-19 pandemic, assistance to PEP cases was slightly lower compared to 2019 (−4%), with a 10% decrease in 2021 compared to 2020^24^. The recommended follow-up routine testing was 30 and 120 days after starting PEP. However, in 2020, with the limitations imposed by the COVID-19 pandemic, a self-test was requested to be carried out in 30 days and a return to the service only in 120 days. Despite this guidance, the 30-day return occurred, showing a greater adherence than the 120-day return. Even before the pandemic, we found that adherence to the 120-day return (12.2%) was very low and worse than in other studies^11^, perhaps due to the fact that an only approximate return date of 30 days was provided and, in case of absence, the user had no other suggested date to return. Even in cases where a later, (e.g. 120 days) return is emphasized, the patient may feel that the 30-day evaluation is sufficient, disregarding further follow-up. In view of this and the possibility of delay in seroconversion, in 2023 we started to orient the first return within 45 days after the start of the PEP (the current limit for the first return according to the Brazilian guideline)^9^ and, if unable to attend, the return within 4 months, both with approximate dates. The UK guideline seems more coherent to this view when considering the delay of a possible seroconversion using antiretrovirals, as it waits at a minimum of 45 days after completion of the PEP course. If the 28-day PEP course is completed, this is 73 days (10.5 weeks) post-exposure^11^.

By evaluating the adherence of those who sought the service and obtained PEP release, we intend to propose a more feasible returns scheme that makes it possible to reduce the loss of opportunities for proper HIV infection diagnosis in these individuals that used PEP, avoiding missed diagnosis due to PEP suppression of biomarkers of infection.

In conclusion, the PEP return protocol in 30 and 120 days did not seem adequate with low adherence at all dates. As the highest adherence is still verified in the first follow-up, very close to the end of the PEP, testing only at this time may increase the chances of false negative results. The second return in 120 days seems very distant from the event, and the user may not return. It is of paramount importance in this scenario to identify a new infection if present and offer proper treatment and consequently break the chain of transmission. We strongly suggest the incorporation of some recommendations of the UK Guideline which suggests that services use local mechanisms, including text/email reminders, to encourage adherence to post-exposure HIV testing^11^. Studies are needed to define a better time that can reconcile test capabilities to detect infection to greater adherence. Strategies to identify infections occurring before or during PEP need to be implemented to avoid discontinuation of a PEP regimen that can be providing viral control and potentially favor future cure strategies.

## Data Availability

All data produced in the present work are contained in the manuscript

